# HIV and the Seroprevalence of SARS-CoV-2

**DOI:** 10.1101/2021.04.29.21256302

**Authors:** Robert L. Stout, Steven J. Rigatti

## Abstract

**Importance:** Healthy HIV-positive insurance applicants have a similar risk of infection and produce a similar antibody response as HIV-negative applicants infected with COVID-19.

**Objective:** Study the seroprevalence and immune response to COVID-19 in healthy HIV-positive life insurance applicants.

**Design:** From January 2020 to March 2021, we examined the seroprevalence of COVID-19 in all HIV-positive applicants. Antibody level in 340 age and sex matched COVID-19 positive applicants, 170 HIV-positive and 170 HIV negative, are compared. The data was de-identified of all personal information and separated by month, age and sex.

**Participants:** Self-reported healthy HIV positive life insurance applicants were tested for antibodies to COVID-19.

## Introduction

Immunodeficiency may place individuals at increased risk of SARS-CoV-2^1,2^ illness. Because COVID-19 infection may be asymptomatic or minimally symptomatic, counts of officially reported cases may substantially underestimate the overall burden of infection^3,4^.

We investigate the seroprevalence of COVID-19 in otherwise healthy HIV-positive life insurance applicants. For HIV-positive applicants to be considered for life insurance, they generally must meet current treatment guidelines, have undetectable viral loads, have been on high-activity antiviral therapy (HAART) for several years, have a CD-4 lymphocyte count greater than 300-500 cell/microliter and have no other significant comorbidities.

## Methods

From January 2020 to March 2021, a national adult convenience sample of self reported healthy life insurance applicants were evaluated for the presence of antibody to HIV with the Roche Elecsys HIV combi PT test^5^.

1,191 HIV-positive samples were tested for total antibody to nucleocapsid protein with the Roche SARS CoV-2 total antibody test. The Roche SARS CoV-2 assay has a reported sensitivity and specificity of 99.5% and 99.8% respectively^6^. All testing was done at Clinical Reference Laboratory, Lenexa, KS.

To investigate if HIV was associated with a detectable difference in immunological response, we examined COVID-19 antibody levels in age and sex-matched HIV positive and negative applicants.

We recorded applicant age, sex, and month tested and deleted all personal data. Statistics were Chi-square and t-test to test differences between positive and negative groups as appropriate with a selected significance level of 99%. All statistical analyses were with R(version3.6.1)^7^ and R-studio (version1.2.1335)^8^.

The study conforms to the recommendations of STROBE for Cross-sectional epidemiology studies, www.equator-network.org.

All participants signed disclosures indicating that results may be used for research purposes. Western IRB (Puyallup, WA) reviewed the study under the Common Rule and applicable guidance and determined it is exempt under 45 CFR § 46.104(d)(4) using de-identified study samples for epidemiologic investigation. WIRB Work Order #1-1324846-1.

## Results

We tested 1,191 HIV-positive applicants for antibodies to COVID-19. Of these, 893 (74.5%) were male, 296 (24.5%) were female with median ages of 40 years (IQR: 33, 40) and 45 years (IQR: 38, 53), respectively. Among the SARS-CoV-2 seropositive cases116 (75.8%) were male with a median age of 40 (IQR: 31,47), and 37 (24.2%) were female with a median age of 44 (IQR: 34.5,52.5). The difference in age and sex between the COVID-19 seronegative and seropositive groups was not significant (p_age_ = 0.23, p_sex_ = 0.54). The overall SARS-CoV-2 seroprevalence in HIV positive females was 12.8% (38 out of 297), and was 12.9% (115 out of 894) in HIV positive men, an insignificant difference.

During the study period, SARS-CoV-2 seroprevalence for HIV-positive individuals increased from 7/81 (8.7%) in May 2020 to 34/103 (33%) in March of 2021(Table 2). COVID-19 antibody levels in the age and sex-matched control plus HIV positive groups varied from 1.1 to > 500 cut-off intensity units (COI), a measure of the amount of antibody present. For age and sex matched COVID positives, the HIV positive population had a higher proportion of individuals with lower COVID antibody levels than HIV negative individuals, but the distributions of antibody levels are not statistically different (Figure 1 and Table 4).

**Table 1:**
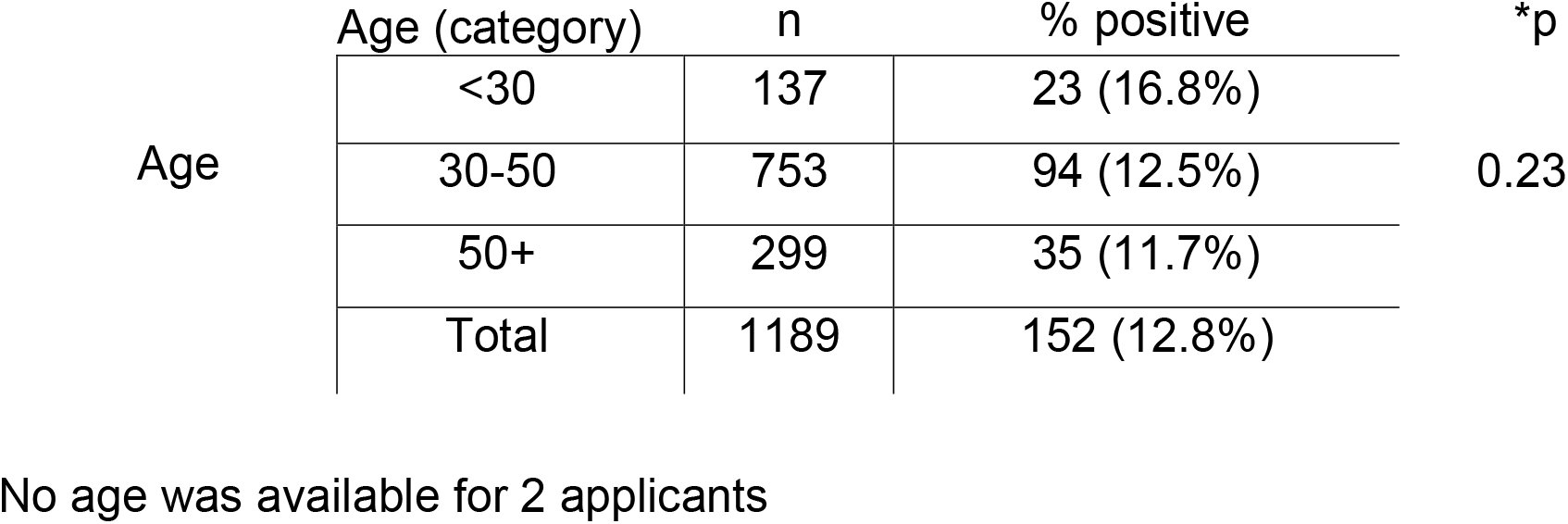
SARS-CoV-2 Seropositivity by Age in the HIV-positive populations.

**Table 2:** COVID-19 Seroprevalence for HIV-positive applicants by sex.

|  |  | Population<br>N (%) | Positive for COVID-19<br>antibodies N (%) | *p |
| --- | --- | --- | --- | --- |
| Sex | Female | 296 (24.2%) | 37 (24.2%) | 0.59 |
|  | Male | 895 (75.8%) | 116 (75.8%) |  |
|  | Total | 1191 (100%) | 153 (9.6% of total) |  |

\*p values based on Pearson Chi-square, 2 tailed test.

**Table 3:**
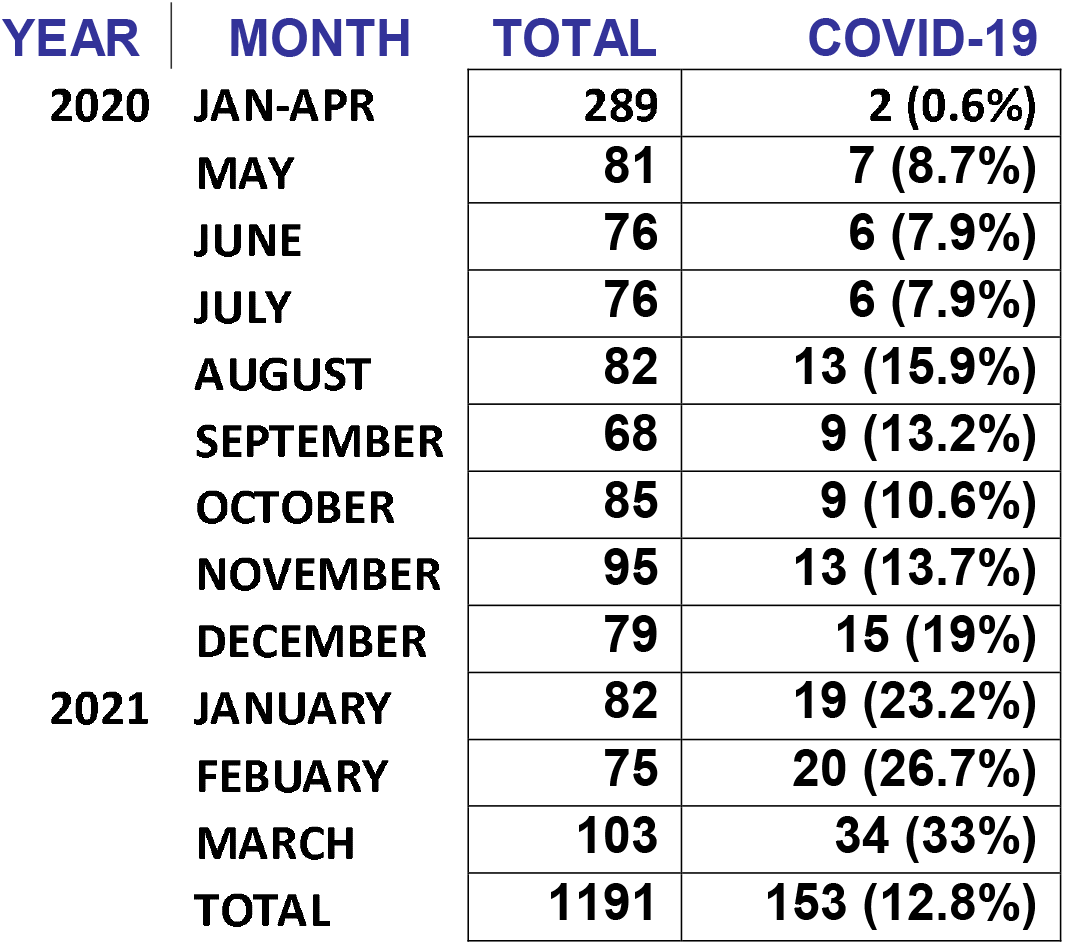
HIV positive samples from January 2020 to March 2021 tested for CVOID-19 antibody.

**Table 4:**
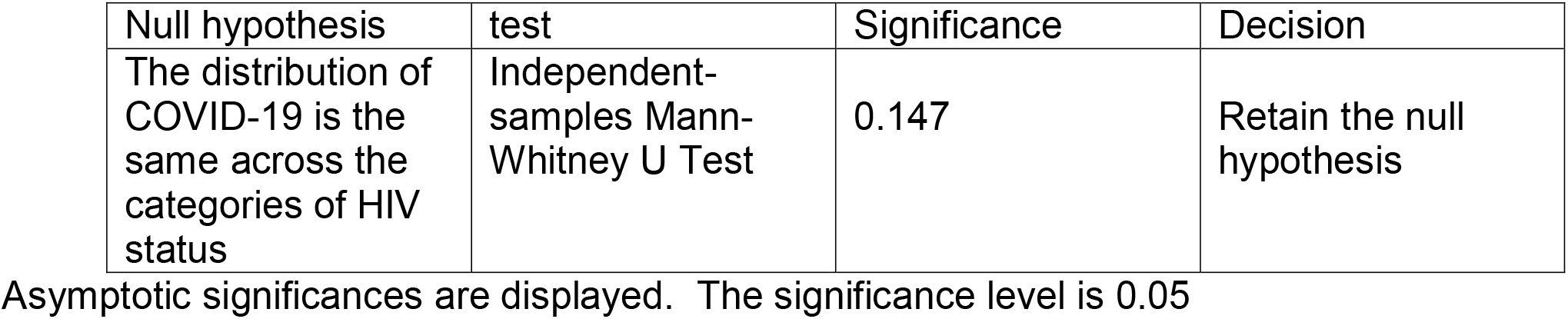
Serum antibody levels to COVID-19 in HIV positive and negative applicants.

| Null hypothesis | test | Significance | Decision |
| --- | --- | --- | --- |
| The distribution of COVID-19 is the same across the categories of HIV status | Independent-samples Mann-Whitney U Test | 0.147 | Retain the null hypothesis |

Asymptotic significances are displayed. The significance level is 0.05

**Figure 1.**
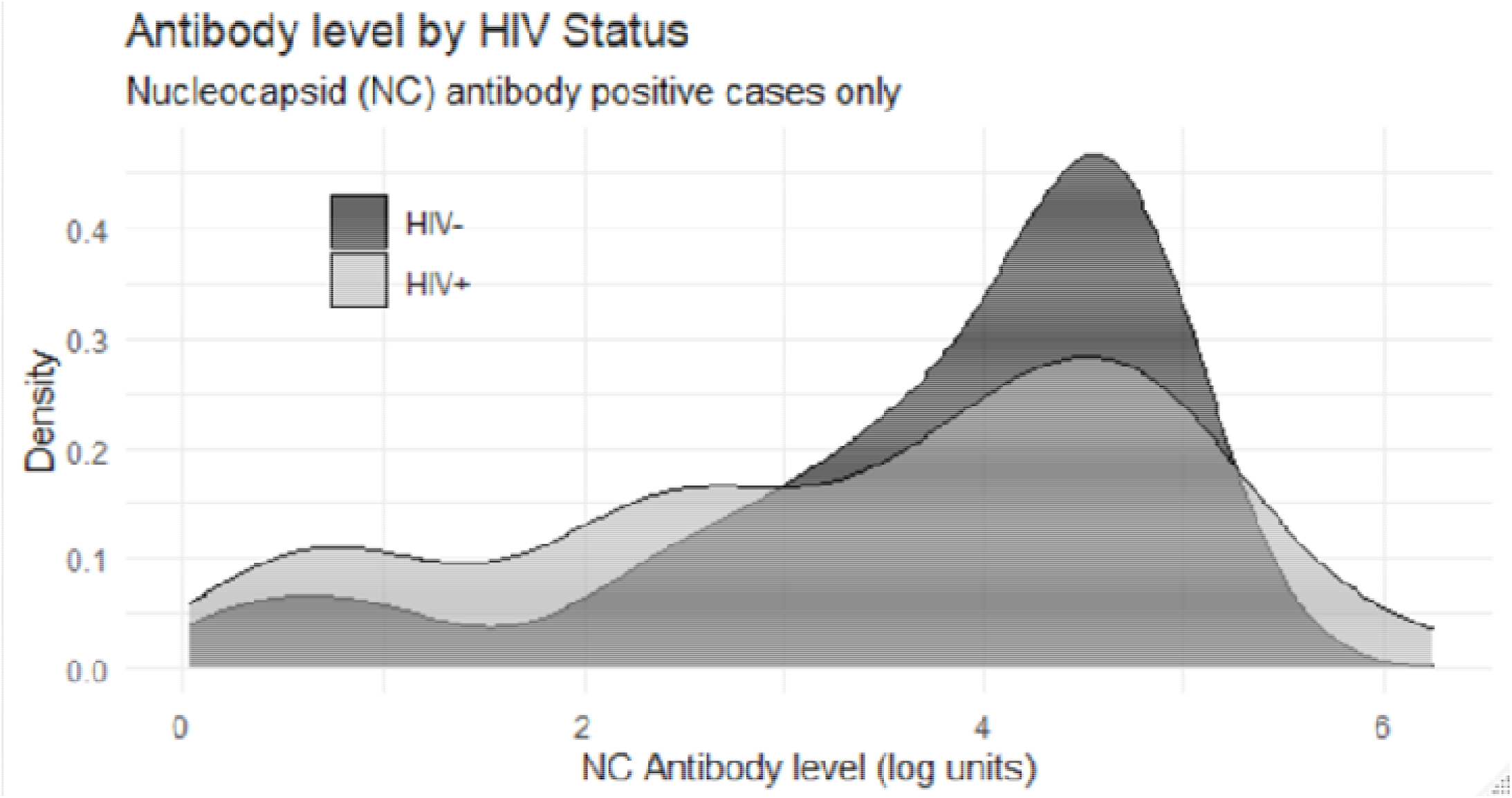
Comparison of COVID-19 antibody levels in HIV positive and negative serum samples.

## Discussion

Previous studies of SARS-CoV-2 infection in the HIV-positive patient population have investigated a possible increase in this population’s adverse outcomes. Richardson *et al* reported on the association of severity of SARS-CoV-2 in 5,700 patients in New York City though there were only 21 HIV-positive cases in the study^9^.

Likewise, Tesoriero *et al* has reported an increase in SARS-CoV-2 severity and death in a population of patients studied in New York^10^. That report includes a larger, more heterogeneous population that suggests a greater severity and mortality risk for HIV patients who become infected with COVID-19.

While public health has consistently warned immunocompromised patients to avoid risk; patient compliance is unknown and poorly studied.

Jérémy Dufloo *et al*^12^ have reported that antibody in both symptomatic and asymptomatic patients is protective. The authors also note that, while the asymptomatic patient may have lower levels of antibody, they are still protective. They report that sera from asymptomatic individuals neutralize the virus, activate Antibody Dependent Cellular Cytotoxicity (ADCC) and trigger complement deposition using replication competent SARS-CoV-2 or reporter cell systems.

## Limitations

Limitations of the study include self-reported health status (well) and possible misrepresentation by the applicant. There was no attempt to determine if the antibody was protective. Even with these limitations the study validates the need for ongoing population-wide surveillance.

## Conclusion

This report is based on a sample from an otherwise healthy HIV-positive population that appear to be compliant with current clinical treatment guidelines. As of March 2021, the seroprevalence of asymptomatic SARS CoV-2 infections in the HIV-positive population is 33%. Anti-COVID-19 antibody levels in HIV positive individuals, in the absence of other comorbidities, suggest that they should have a clinical course similar to HIV negative individuals. The high seroprevalence of COVID-19 antibody in HIV patients suggests that the current public health mitigation strategy may be inadequate to reduce the asymptomatic spread of infection or that the public is failing to fully comply with those recommendations.

## Data Availability

Data are available to non-commercial organizations for research use.

## Acknowledgement

Both authors had full access to all the data in the study and take responsibility for the integrity of the data and the accuracy of the data analysis.

## Statement of author contribution

Robert Stout directed the testing of samples, combined the demographic data and then deleted all identifiable personal information. Reviewed all patient data for completeness and prepared the initial prevalence estimates. Steve Rigatti analyzed the prevalence for the data set and prepared the statistically analysis. Both authors prepared the text.

